# Neurofilament light chain reference values in serum and cerebrospinal fluid: a bi-compartmental analysis in neurological diseases

**DOI:** 10.1101/2024.11.21.24317697

**Authors:** Steffen Halbgebauer, Veronika Klose, Badrieh Fazeli, Paula Klassen, Christoforos Alexudis, Gabriele Nagel, Angela Rosenbohm, Dietrich Rothenbacher, Nerea Gomez de San Jose, Simon Witzel, Zeynep Elmas, Maximilian Wiesenfarth, Joachim Schuster, Johannes Dorst, Andre Huss, Franziska Bachhuber, Markus Otto, G. Bernhard Landwehrmeyer, Albert C. Ludolph, Hayrettin Tumani, the ALS Registry Swabia study group

## Abstract

**Background:** Concentrations of neurofilament light chain (NfL), a neuroaxonal damage marker, increase with age. Therefore, age-dependent reference values are important in clinical practice. However, so far these have only been established with a bead based system and age-dependent z-scores for CSF are missing. In addition, we here propose how the combined analysis of CSF and serum NfL could help in the discrimination between central (CNS) and peripheral nervous system (PNS) axonal degeneration.

**Methods:** For the calculation of age-reference values for serum and CSF 1514 NfL concentrations of control subjects determined with the microfluidic Ella system, were applied.

**Results:** Age-dependent NfL levels were calculated with additive quantile regression and presented with percentiles and z-scores. We observed a non-linear increase of NfL in serum and CSF. The spearman r of the association with age was 0.81 (95% CI: 0.78-0.83), p<0.0001 and 0.82 (95% CI: 0.79-0.85), p<0.0001 for serum and CSF NfL, respectively. Serum and CSF NfL levels were also associated with each other (r=0.68 (95%CI: 0.62-0.73), p<0.0001). Furthermore, we used this association to establish a bi-compartmental CSF and serum NfL model allowing to differentiate between peripheral or central origin of neurodegeneration.

**Conclusions:** The age-reference curves corroborate findings of an exponential elevation of NfL in serum and CSF with increasing age. As NfL values from different platforms are not interchangeable this is of additional high relevance. Moreover, the association between CSF and serum NfL values could be applied for clinical use regarding overlapping symptoms of CNS and PNS based neurological diseases.

**What is already known on this topic:** Neurofilament light chain levels in cerebrospinal fluid (CSF) and blood are widely accepted neuroaxonal damage markers which strongly correlate with age.

**What this study adds:** Age-dependent reference values and z-scores for both serum and CSF for the neurofilament light chain protein (NfL) using the microfluidic ELLA analysis platform. Novel approach for the discrimination between central and peripheral axonal damage using a bi-compartmental NfL model.

**How this study might affect research, practice or policy:** Easier and age-corrected interpretation of NfL values in clinical practice. Possibility to distinguish central from peripheral damage by analyzing both blood and CSF NfL.

## Introduction

Neurofilaments represent the principal cytoskeletal component of neurons, providing structural stability through the formation of fibrillary networks. They are highly abundant in neuronal axons, where they are involved in a number of tasks, including axonal transport and dendritic branching [1, 2]. The most abundant neurofilament is the neurofilament light chain (NfL), a 68 kDa intermediate filament protein. NfL is also the neurofilament that has been the subject of the most extensive investigation to date, serving as a fluid biomarker in neurological diseases [3]. Following neuroaxonal damage, NfL is released into the CSF and subsequently drained into the bloodstream, where it can be quantified using state-of-the-art technology such as microfluidic assays (Ella) or single molecule array (Simoa). Given its expression pattern, NfL levels are markedly elevated in motor neuron diseases such as amyotrophic lateral sclerosis (ALS) [4–6], but increased levels can also be observed in numerous other conditions, including multiple sclerosis [7, 8] and frontotemporal dementia [9, 10]. This offers the opportunity to utilize NfL levels in CSF and blood as diagnostic, progression and monitoring markers in the clinic and clinical trials. However, it should be noted that NfL levels can be affected by comorbidities (e.g. cardiovascular diseases, diabetes mellitus), confounding factors such as BMI or the glomerular filtration rate and, in particular, by age [11, 12]. A number of studies have been conducted to establish age-related reference NfL levels in control patients with a wide age range [13–15]. These studies employed different versions of the Simoa NfL kit, although other platforms have also been used in studies on NfL. One such platform is the microfluidic Ella NfL test, which has been shown to correlate well with the Simoa assays [16–18]. The Ella assay is applicable in daily clinical routine and utilizes the same antibodies as the Simoa kits. However, Simoa and Ella NfL kits demonstrate disparate absolute NfL concentrations. Consequently, the objective of this study was to establish age-related NfL reference values in serum and CSF using the microfluidic Ella NfL technology in subjects without neurodegenerative and neuroinflammatory diseases.

Furthermore, we set up z-score references for serum and CSF NfL levels which can be applied across platforms. Our study covers a wide age range from 18-89 years (50.3% female) using in total 1514 NfL values for the age reference NfL concentration calculations. In addition, from approximately 400 control patients CSF and serum was available which we used to generate a bi-compartmental model to help distinguish between peripheral and central origins of neuroaxonal damage. To validate the bi-compartmental model we additionally analyzed CSF and serum NfL levels in four neurological disorders.

## Material and Methods

### Patients

In this study, we used NfL values from control subjects from two different studies performed at Ulm University Hospital. The first cohort consisted of patients ranging from age 33-89 enrolled in the population-based ALS Swabian registry (ethics votes No. 11/10, No. B-F-2010-062 and No. 7/11300) as control cases (n=576). The study has been previously described [19–22]. All members of the registry are listed in the supplementary materials. From this cohort only serum NfL was available.

The second cohort comprised 72 patients with serum and CSF NfL concentrations available, ranging from age 21-88, which were used as control patients in a neurofilament NfL and NfH validation study [4]. To increase the number of samples we additionally analyzed serum NfL in 385 and CSF NfL in another 403 patients from age 18 to 89, which were seen at the Department of Neurology at Ulm University Hospital between 2014-2023 and collected through convenience sampling. These patients did not show clinical or radiological signs for neurodegeneration and an acute inflammation of the central nervous system was ruled out by CSF analysis (diagnoses can be found in table S1). A flow chart of the patient selection can be found in the supplementary materials (Fig. S1).

For the evaluation of the bi-compartmental model, we additionally measured NfL from patients with a neurological disorder seen at the University Hospital Ulm (Fig. S1). The multiple sclerosis (MS) patients were diagnosed with progressive MS according to the revised McDonald criteria [23]. For the diagnosis of Guillain–Barré syndrome (GBS) the consensus guidelines according to Leonhard et al. 2019 [24] and for idiopathic intracranial hypertension (IIH) according to Mollan et al. 2018 [25] were applied. CSF and serum NfL levels of 10 MS, 9 GBS and 12 IIH patients were analyzed for this study. CSF and serum NfL concentrations of 10 amyotrophic lateral sclerosis (ALS) patients were taken from a previously published study where ALS was diagnosed according the El Escorial criteria [4, 26]. Control and disease patients seen at the neurological department at the University Hospital Ulm suffering from acute or chronic renal insufficiency were excluded from the study. The examination was approved by the local ethics committee (approval number Ulm 20/10) and conducted following the Declaration of Helsinki. All participants gave their written informed consent to participate in the study.

### Sampling and NfL measurement

CSF and blood samples were collected by lumbar puncture and venous sampling, respectively. Samples were centrifuged at 2000 g for 10 min and CSF supernatant and the extracted serum aliquoted and frozen within 30min at -80°C. Both CSF and serum samples were stored in polypropylene tubes.

For NfL quantification in CSF and serum the microfluidic Ella platform (BioTechne, Minneapolis, USA) was used. The analyses were performed according to the manufacturer’s instructions.

### Statistics

For the analysis of the effect of age on NfL levels due to a nonlinear relation an additive quantile regression analysis was performed based on the control population using RStudio (Version 4.3.1). For visualization Python (Version 3.11.5) was applied. According to this analysis we calculated the z-sores for the control group. Moreover, we applied the outcome of the model to determine the z-scores of the disease cohorts.

GraphPad Prism V.10.3.1 (GraphPad, Software, La Jolla, California, USA) was used to calculate the association between serum and CSF NfL applying the spearman rank correlation and for the comparison between the diagnostic groups using Kruskall-Wallis followed by Dunn’s post hoc test. For female and male GFAP level differences between age groups the Mann-Whitney U test was applied. A p<0.05 was indicative of statistically significant results.

## Results

### NfL reference concentrations by age

A total of 1514 NfL concentrations were utilized for the establishment of reference values by age. The median NfL concentrations for the serum and CSF control cohort, along with the demographic parameters, can be found in Table 1.

**Table 1:** Demographic and NfL parameters of the analyzed cohort.

|  | Control Patients for Serum NfL | Control Patients for CSF NfL |
| --- | --- | --- |
| N | 1033 | 475 |
| Age (years) mean±SD | 56±18 | 45±19 |
| f/m | 520/513 | 302/173 |
| Serum NfL [pg/ml]<br>Median (IQR) | 19 (10-27) | n/a |
| CSF NfL [pg/ml]<br>Median (IQR) | n/a | 359 (218-589) |

CSF, cerebrospinal fluid; f, female; m, male; NfL, neurofilament light chain

Figure 1A depicts the individual serum NfL values for patients aged 18 to 89, as well as the calculated regression lines for the median and the 25th, 80th, and 95th percentiles. The serum NfL values demonstrate an increase with age, with a median (interquartile range) of 5.5 pg/ml (4.0-7.0 pg/ml) in the group of patients below 25 years of age and 42.5 pg/ml (29.5-63.5 pg/ml) in patients above 80 years of age. The correlation between age and serum NfL levels was found to be strong, with a Spearman r of 0.81 (95% CI: 0.78-0.83), p<0.0001. Figure 1B displays the z-scores for ranges from 0 to 2, rather than percentiles. The effect of the body mass index and sex on NfL levels can be found in the supplementary materials.

**Figure 1:**
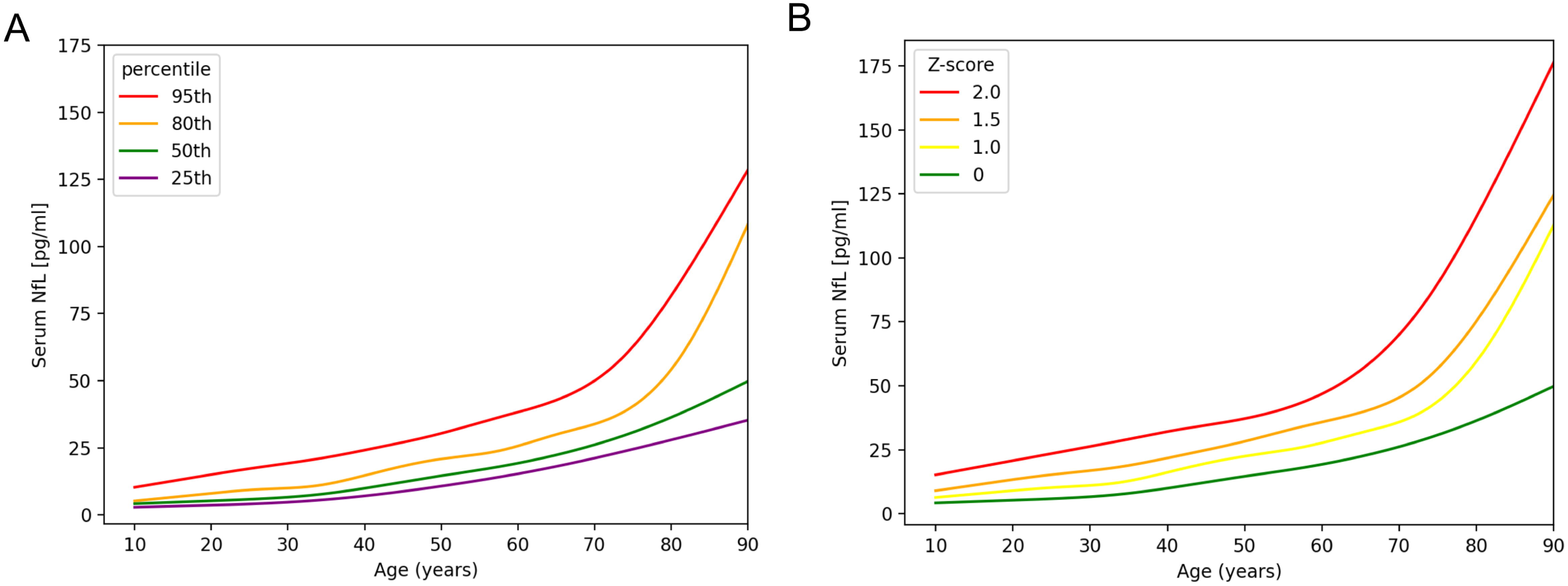
Serum NfL age-reference curves. **A** depicts the serum NfL levels as percentiles from 25 to 95. Serum NfL concentrations strongly increase over time. **B** displays the serum NfL concentrations as z-scores ranging from 0-2. For modelling the NfL concentration with age additive quantile regression was applied. Abbreviations: NfL, neurofilament light chain.

Figure 2 displays CSF NfL levels which increase from 172 (148-210) pg/ml in the group aged below 25 to 1322 (1084-1820) pg/ml in the age group above 80. Figure 2A shows the percentiles, while Figure 2B shows the CSF NfL z-scores ranging from 0-2. As serum NfL levels correlate with age, CSF NfL concentrations are also associated, with an r of 0.82 (95% CI: 0.79-0.85), p<0.0001.

**Figure 2:**
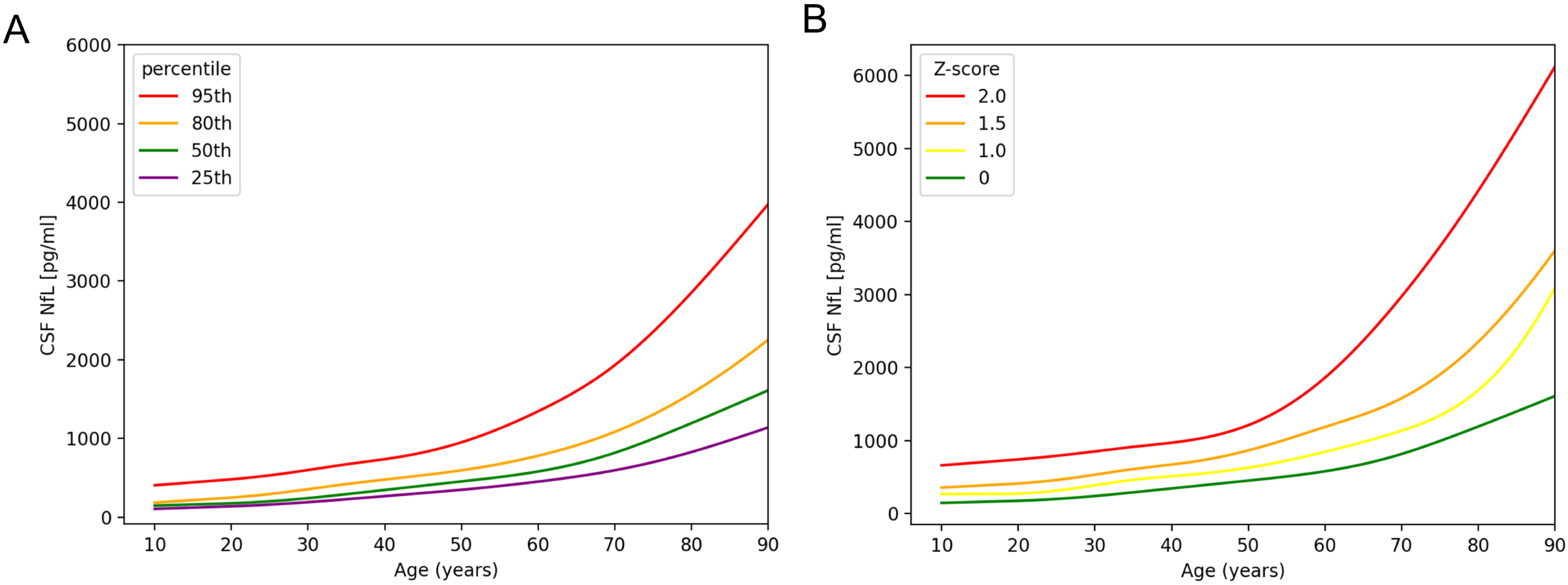
CSF NfL age-reference curves. **A** depicts the CSF NfL levels as percentiles from 25 to 95. The CSF NfL levels demonstrate a clear increase with age. **B** displays the CSF NfL concentrations as z-scores ranging from 0-2. For modelling the NfL concentration with age additive quantile regression was applied. Abbreviations: CSF, cerebrospinal fluid; NfL, neurofilament light chain.

The serum and CSF NfL median and 95^th^ percentiles for the respective age groups are displayed in table 2. Z-score values can be found in the supplementary materials.

**Table 2:** Age-specific 50% and 95% NfL percentiles in serum and CSF in control patients:

| Age [years] | Serum 50th percentile [pg/ml] | Serum 95th percentile [pg/ml] | CSF 50th percentile [pg/ml] | CSF 95th percentile [pg/ml] |
| --- | --- | --- | --- | --- |
| 20 | 5 | 15 | 175 | 479 |
| 25 | 6 | 17 | 200 | 530 |
| 30 | 7 | 19 | 240 | 598 |
| 35 | 8 | 21 | 291 | 669 |
| 40 | 10 | 24 | 344 | 736 |
| 45 | 12 | 27 | 398 | 821 |
| 50 | 15 | 30 | 452 | 951 |
| 55 | 17 | 34 | 508 | 1126 |
| 60 | 19 | 38 | 578 | 1343 |
| 65 | 22 | 43 | 678 | 1605 |
| 70 | 26 | 50 | 816 | 1933 |
| 75 | 31 | 63 | 995 | 2359 |
| 80 | 36 | 82 | 1192 | 2851 |
| 85 | 43 | 105 | 1400 | 3400 |

Abbreviations: CSF, cerebrospinal fluid; NfL, neurofilament light chain.

A total of 384 patients had CSF and serum NfL values available for analysis in a bi-compartmental model. As illustrated in Figure 3A, the correlation analysis between serum and CSF NfL values revealed a medium to strong correlation (r = 0.67 (95% CI: 0.60-0.72), p < 0.0001). The letters A-D define different areas indicative of not elevated (A), elevated CSF and serum (B), only elevated serum (C) and only elevated CSF (D) NfL values compared to the control cohort. In a next step, we employed the bi-compartmental model from figure 3A in a cohort of different neurological diseases. Four patient cohorts consisting of MS, ALS, GBS and IIH were analyzed for absolute CSF and serum NfL values and z-scores (Fig. S2). The boxplots in figure S3 in the supplementary materials depict that the CSF to serum ratio of NfL is similar for MS and ALS but seems to be lower for GBS and higher for IIH.

**Figure 3:**
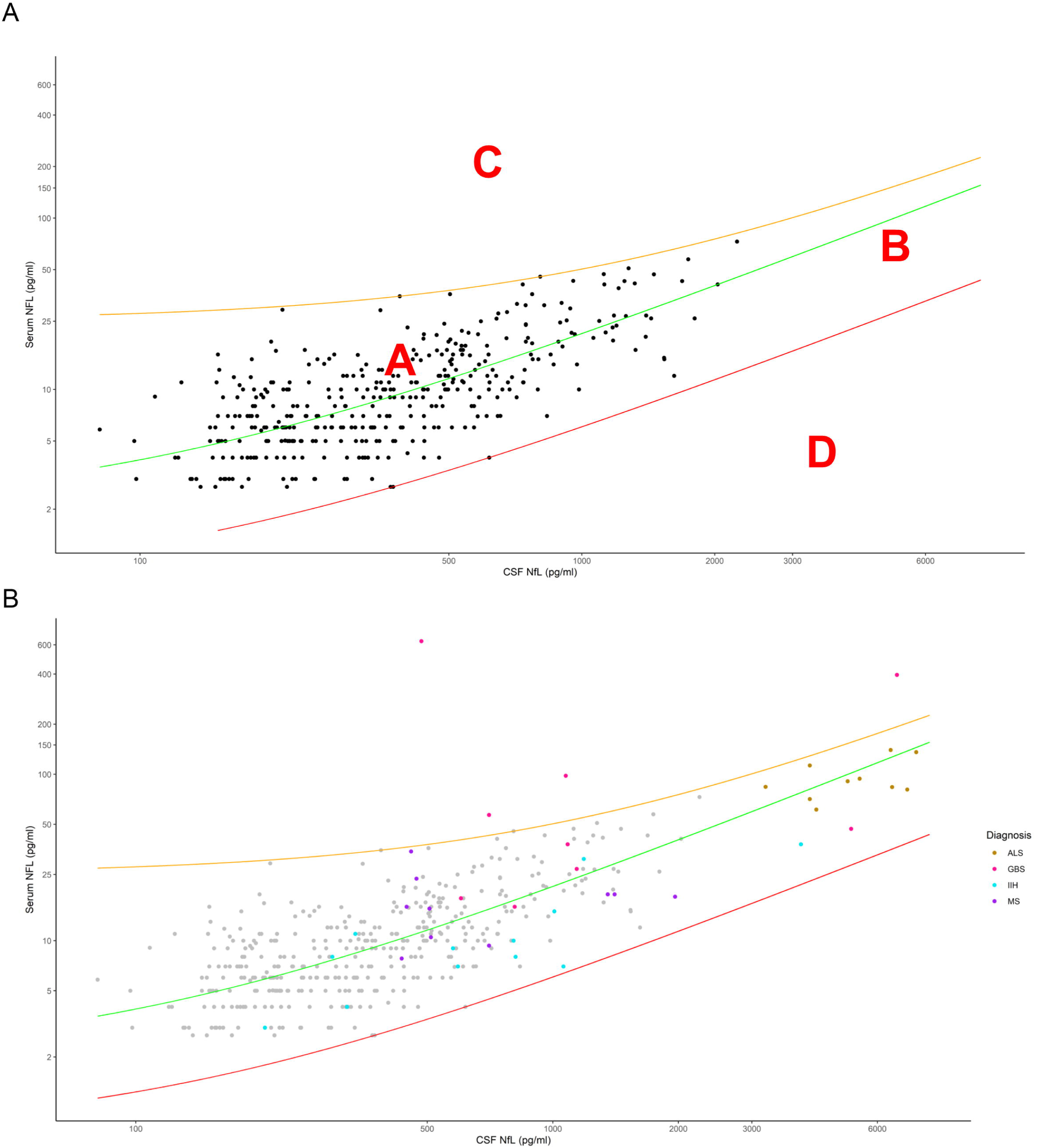
CSF versus serum NfL. Figure (**A)** displays the corresponding CSF and serum NfL values of 384 control patients. In green the median simple linear regression line is shown. Orange and red lines depict the corresponding +3SD and -3SD lines. The letters A-D define areas indicative of (A) normal, (B) increased CSF and serum, (C) only or predominantly increased serum and (D) only increased CSF NfL values. The orange and red lines represent the +3SD and -3SD regression lines. In (**B)** the same graph is illustrated including patients of four disease cohorts (ALS, MS, GBS and IIH). Abbreviations: ALS, amyotrophic lateral sclerosis; CSF, cerebrospinal fluid; GBS, Guillain-Barré-Syndrom; IIH, Idiopathic intracranial hypertension; NfL neurofilament light chain; MS, multiple sclerosis; SD, standard deviation.

In Figure 3B, the four disease groups were illustrated in the generated graph from figure 3A. The MS patient group can be found in area A and the ALS group in B. GBS patients depicted high serum levels compared to CSF and can therefore partly be seen in area C. The IIH patient cohort depicted the highest CSF to serum ratio and are consequently below the median regression line. However, no patient could be detected in area D.

The mean ratio of CSF and serum NfL was determined to be 44. Figure S4 in the supplements depicts the quotient of CSF and serum NfL dependent on age. With an r of 0.01 (95%CI -0.1-0.1, p=0.9) we found no correlation.

## Discussion

The present study for the first time describes NfL values in serum and CSF depending on age analyzed by microfluidic Ella analysis. Our results confirm the current literature and age NfL reference values analyzed with Simoa as both serum and CSF NfL concentrations increase with age [13–15]. Moreover, our findings support the notion that the correlation between age and NfL elevation is not linear [13, 15].

We found a prominent elevation of NfL values above the age of 80 for both serum and CSF NfL. This effect was found to be quite strong and might be in addition to normal aging be attributed to underlying presymptomatic pathological changes which were not yet detectable in clinical routine. Nevertheless, this finding is significant as it underscores the necessity for caution when interpreting NfL levels in individuals beyond the age of 80.

NfL z-scores indicate the number of standard deviations a single value is above the mean of all measured control patients. In comparison to absolute values, z-scores exhibit several advantages. They are normally distributed, can also have negative values and are independent of the platform applied to measure the NfL concentrations. Our findings display a similar blood z-score pattern as published by Benkert and colleagues [15]. Furthermore, we calculated z-scores for CSF NfL, which were not previously available in the literature.

Our correlation analysis of nearly 400 samples revealed a strong association between CSF and serum NfL values. This corroborates data from a meta-analysis on the correlation of blood and CSF NfL, which found a nearly identical r value [27]. However, the correlation is weaker than in our recent NfL study [4], where neurodegenerative diseases, especially ALS, were included in the correlation analysis. Nevertheless, the graphical illustration of the association of CSF and serum NfL concentrations in control patients could be beneficial for the interpretation of patient NfL values in the clinic. If CSF and blood NfL patient values are available, the clinician will be able to compare the plotted values to those of the controls as depicted in our bi-compartmental model. This would provide further insight into whether a possible elevation in blood is CNS or PNS derived, depending in which area (A-D) the value is located. Area A represents normal NfL levels. In addition to the control, in our study the MS patients are found in the upper part of this area which corroborates data of NfL in MS being slightly increased compared to healthy individuals in the same age range but only reach really high levels during an active relapse [28]. Area B depicts increased NfL levels in CSF and blood, which is highly indicative of a CNS-derived neurodegeneration. The ALS cohort as a classical CNS-derived disease group could be detected in this area confirming the hypothesis. Area C represents predominantly increased levels in blood which could be indicative of a more peripheral origin of axonal damage as seen in some neuropathies [29, 30]. Some patients of the GBS cohort could be detected in the area which is highly suspicious of a strong peripheral neuroaxonal damage for these patients. The model thereby was also able to help in the differential diagnosis to ALS. Area D, which exhibits elevated levels exclusively in CSF, is the most challenging to interpret as CSF NfL levels should also reach the bloodstream. A possible explanation could be an impaired CSF outflow as demonstrated by Engel and colleagues [31]. However, our cohort of IIH patients which also depicted a high CSF to serum NfL ratio as seen by the colleagues [31], could also not be detected in this area. It remains to be seen if any disease group can be attributed to this area. Furthermore, it should be noted that the bi-compartmental graph is a preliminary model and in order to clearly define the areas A-D, a much larger sample size is required than was available for this study.

The average CSF NfL levels were found to be 44 times higher than those in blood, consistent with Simoa results, which detected levels approximately 42 times higher in CSF than in blood [8]. In contrast to CSF or serum NfL values alone, the ratio of both does not appear to be associated with age, indicating that the outflow of NfL from the CSF into the bloodstream may be independent of age.

The present study contributes to the existing literature as it is highly important to validate NfL data so far mostly only generated with the Simoa assay, also using other platforms. In addition, it is crucial for Ella NfL users to compare their NfL concentrations to reference values generated with the same platform as the absolute values of Simoa and Ella are not interchangeable [16–18] and harmonization projects with NfL reference materials are under way but are far from being implemented [32].

It should be noted, that the present study is not without limitations. Firstly, the CSF control cohort is smaller than the serum cohort. Secondly, in the age group above 80, there is a need for further analysis of a larger number of patients.

In conclusion, the age-specific reference NfL percentiles for serum and CSF can be applied to compare analyzed patient data in the clinic. Furthermore, NfL z-scores could be especially valuable in the follow-up measurements of patients, for instance under treatment. Additionally, the simultaneous measurement of NfL in CSF and blood might assist in the identification of the origin of axonal damage and help in the differential diagnosis of neurological diseases.

## Data Availability

All data produced in the present study are available upon reasonable request to the corresponding author.

## ACKNOWLEDGEMENTS

We thank the Ilona Kraft-Overbeck, Ines Dobias and Nicola Lämmle for their excellent field work, and Gertrud Feike, Sarah Enderle and Birgit Och for their excellent data management and technical support as well as all patients and healthy controls for participation in the study.

## AUTHOR CONTRIBUTIONS

All authors made substantial contributions to conception and design, and/or acquisition of data, and/or analysis and interpretation of data.

All authors gave final approval of the version to be submitted and agree to be accountable for all aspects of the work in ensuring that questions related to the accuracy or integrity of any part of the work are appropriately investigated and resolved.

Conception and design of the study: SH, HT; Sample collection and data management: SH, VK, BF, PK, CA, GN, AR, DR, NG, SW, ZE, MW, JS, JD, AH, FB, MO, GBL, ACL, HT; Study management and coordination: SH, HT; Statistical methods and analysis: SH, VK, HT; Interpretation of results: SH, VK, BF, MO, ACL, GBL, HT; Manuscript writing (first draft): SH, HT; Critical revision of the manuscript: SH, VK, BF, PK, CA, GN, AR, DR, NG, SW, ZE, MW, JS, JD, AH, FB, MO, GBL, ACL, HT.

