## Supplement for "Neurofilament light chain reference values in serum and cerebrospinal fluid: a bi-compartmental analysis in neurological diseases"

Dr. Felix Geser, Christophsbad Göppingen

Dr.med. Karsten Henkel, Gerontopsychiatrie Christophsbad Göppingen

Dr. med. Martin Zinkler, Kliniken Heidenheim

Dr.med. Ralf Kozian, Gerontopsych.Vinzenz von Paul Hospital, Rottweil

Prof. Dr. med. Dr. phil. Martin Bürgy, Allgemeinpsych Zentrum f. Seelische Gesundheit, Klinikum Stuttgart, Bad Cannstatt

Priv.-Doz. Dr. med. Christine Thomas, Gerontopsych Zentrum f. Seelische Gesundheit,Klinikum Stuttgart, Bad Cannstatt

Dr.med. Stefan Spannhorst, Zentrum für Seelische Gesundheit,Klinikum Stuttgart, Bad Cannstatt

Priv.-Doz. Dr. med. Matthias Munk Unilklinik Tübingen

Prof.Dr. Christoph Laske Uniklinik Tübingen

PD Dr.med.Daniel Schüpbach ZfP Weinsberg, Klinikum am Weissenhof, Allgemeinpsych West

PD Dr.med.Heinz Grunze ZfP Weinsberg, Klinikum am Weissenhof, Allgemeinpsych Ost

Dr. Rainer Schaub, Gerontopsych ZfP Weinsberg, Klinikum am Weissenhof

Dr. Jochen Gebhardt, Gerontopsychiatrie ZfP Wiesloch

Dr.med. Hubertus Friederich, Gerontopsych. ZFP Zwiefalten

Dr. Matthias Köhler, Gerontopsych. ZFP Zwiefalten

Dr.med.Alex Gogolkiewicz, Allgemeinpsych.ZfP  Zwiefalten

Prof.Dr. med. Andreas Joos Kliniken Schmieder Gailingen

Prof. Dr. med. Max Schmauß BKH Augsburg

Dr.med.Jessica Baumgärtner BKH Augsburg

Prof. Riepe, Gerontopsych.BKH Günzburg , Geontopsych

Prof. Becker, Allgemeinpsych.BKH Günzburg,  Psych.II

Dr. Andreas Küthmann Bezirkskrankenhaus Memmingen

Dr. Raimund Steber Bezirkskrankenhaus Memmingen

Dr. Ivona Gecui Bezirkskrankenhaus Memmingen

Univ. Prof. Dr. med. Elmar Etzersdorfer Furtbachkrankenhaus, Stuttgart

Dr.med Alexandros Michaelides, Furtbachkrankenhaus, Stuttgart

Prof. Dr. med. Bernhard Connemann, Uniklinik Ulm, Psych. III

Andreas Raether, Gerontopsychiatrie, ZfP Winnenden

### Tables

Table S1: Diagnoses of control patients selected at the Department of Neurology at the University Hospital Ulm

| Diagnosis | Number |
|---|---|
| Acoustic hallucinations | 1 |
| Affection of the facial nerve on the right side | 1 |
| Allergic reaction | 1 |
| Analgesic-induced headache | 1 |
| Arteriitis temporalis | 1 |
| Barbiturate intoxication | 1 |
| Bladder and rectal dysfunction (psychogenic) | 1 |
| Bladder atony of unknown cause | 1 |
| Catatonic stupor | 1 |
| Central retinal vein occlusion right eye | 1 |
| Central visual loss with subsequent headaches (migraine attack) | 1 |
| Cervical spine disc prolapse | 1 |
| Cervical spine syndrome with cervical myalgia (excluded meningitis) | 1 |
| Cervical spine syndrome with disc protrusion | 1 |
| Chron. Cervical spine pain syndrome | 1 |
| Chronic cervicalgia with pseudoradicular radiation | 1 |
| Chronic left-sided lumbalgia | 1 |
| Chronic myalgic syndrome, unclear cause | 1 |
| Chronic tension headache | 1 |
| Clinical gait uncertainty | 1 |
| Coxarthrosis right | 1 |
| Craniomandibular dysfunction left | 1 |
| Deregulated insulin-dependent diabetes mellitus | 1 |
| Developmental Venous Anomaly subcortical | 1 |
| Discrete ptosis on the left | 1 |
| Distal myopathy of the lower extremity | 1 |
| Distal symmetric axonal sensorimotor polyneuropathie | 1 |
| Double images | 1 |
| Drug-induced cephalgia | 1 |
| Exclusion diagnosis of ocular myasthenia | 1 |
| Exclusion of an inflammatory CNS disease with numbness of the right hand | 1 |
| Exclusion of an organic cause for left-sided facial discomfort | 1 |
| Exclusion of inflammatory CNS disease with paraesthesia and hypaesthesia | 1 |
| Exclusion of inflammatory genesis of diffuse tingling paraesthesia | 1 |
| Exclusion of multiple sclerosis | 1 |
| Exclusion of spondylodiscitis | 1 |
| Exclusion of trigeminal nerve affection | 1 |
| Exercise-induced myalgias (muscle biopsy no myopathy) | 1 |
| First migraine attack with aurea | 1 |
| Hemicrania continua with left-sided headache and Horner's syndrome left | 1 |
| Hemiplegic migraine | 1 |
| herpes zoster ophtalmicus | 1 |
| hip contusion | 1 |

| | |
|---|---|
| Hypaesthesia of the right arm of unclear aetiology | 1 |
| Hypaesthesia of the right leg of dissociative origin | 1 |
| Hypaesthesia V2 and V3 left side of the face, exclusion of chronic inflammation | 1 |
| Hypothyroidism | 1 |
| Idiopathic abducens palsy on the right | 1 |
| Idiopathic left abducens paresis | 1 |
| Idiopathic sudden deafness on the right | 1 |
| Impingement syndrome shoulder joint | 1 |
| Intermittent brachial and facial hypaesthesia without evidence of an organic cause | 1 |
| Left frontal headache, excluding secondary cause of headache | 1 |
| left-sided central sensorimotor hemisymptomatics | 1 |
| Lumbago | 1 |
| lupus erythematosus | 1 |
| Medication-induced headache, dizzines | 1 |
| Menstrual migraine without aurea | 1 |
| Migrain with visual aurea | 1 |
| Migraine with temporary hemiparesis | 1 |
| Migraine with vestibular aura | 1 |
| Migraine with visual aurea | 1 |
| Monarthritis right knee | 1 |
| Motor arm plexus palsy | 1 |
| Multifactorial gait disorder | 1 |
| Neuralgiform headache | 1 |
| Neuropathic pain syndrome | 1 |
| No evidence of meningitis | 1 |
| No evidence of neuroborreliosis, erythema migrans after tick bite | 1 |
| Non-specific multilocular tingling paraesthesia | 1 |
| Non-specific paraesthesia of all extremities | 1 |
| Non-specific sensory disturbance of the right arm | 1 |
| Non-specific, fluctuating tingling paraesthesia | 1 |
| Occipital neuralgia | 1 |
| Opressive headaches | 1 |
| Orthostatic vertigo | 1 |
| OSG supination trauma | 1 |
| Pain in right hand of unknown origin | 1 |
| Paraesthesia of the hands and feet on both sides | 1 |
| Paraesthesia of the richt hand | 1 |
| Paraesthesia of the right side of the body | 1 |
| Parainfectious headache, Covid infection | 1 |
| Parese des N. peronaeus profundu | 1 |
| Partial rupture of the popliteus tendon at the femoral origin | 1 |
| Pressure lesion of the lower brachial plexus on the left | 1 |
| Prolong migraine with aura | 1 |
| Proximally emphasised arm paresis | 1 |
| Recurrent left-sided brachial and facial paresthesias | 1 |
| Recurrent polyradiculopathies of the left upper extremity | 1 |
| Recurrent presyncope | 1 |
| Regredient vertigo and tingling paraesthesia of both legs | 1 |
| Residual complaints metatarsophalangeal joint on the right | 1 |
| rheumatoid arthritis | 1 |

| | |
|---|---|
| Right abducens nerve palsy | 1 |
| Right-sided tingling paraesthesia | 1 |
| S1 syndrome left | 1 |
| Sensible C6-syndrom | 1 |
| Sensitive L5 syndrome on the right side | 1 |
| Sinusitis | 1 |
| Spinal canal stenosis HWK 5/6 with myelopathic signal | 1 |
| Stress reaction | 1 |
| Subjective gait disturbance, no evidence of organic genesis | 1 |
| Subjective left sensorimotor hemisymptomatics, exclusion of central genesis | 1 |
| Suspected presyncope | 1 |
| Suspicion of disc prolapse | 1 |
| Symptomatic facial pain on the left of dentogenic origin | 1 |
| Thoracalgia | 1 |
| Tic disorder of unclear aetiology | 1 |
| Tolosa hunt syndrome | 1 |
| Transient Double images | 1 |
| Transient Hemihypaesthesia | 1 |
| Transient hypaesthesia extremities left | 1 |
| Transient Tingling parasthesias | 1 |
| Transient vertigo | 1 |
| Unsystematic feelings of tingling and numbness | 1 |
| Unsystematic sensory disturbance on the left elbow and the soles of the feet | 1 |
| Unsystematic sensory disturbances of unclear cause | 1 |
| ventricular extrasystole | 1 |
| Vertigo of peripheral vestibular origin | 1 |
| Visual impairment left | 1 |
| amnesic aphasia | 1 |
| Adjustment disorder | 2 |
| Anxiety disorder | 2 |
| Benign fasciculations | 2 |
| brachial plexus neuritis | 2 |
| Choreatic hyperkinesia | 2 |
| Cluster headache | 2 |
| Exclusion of a neuromuscular disease | 2 |
| Exclusion of neuroborreliosis | 2 |
| giant cell arteritis | 2 |
| Lumbalgia | 2 |
| Lumboischalgia | 2 |
| Mononeuritis multiplex | 2 |
| Nerve root affection | 2 |
| Oculomotor nerve palsy | 2 |
| Pain disorder | 2 |
| Pansinusitis | 2 |
| Papilledema on both sides | 2 |
| Post-puncture headache, tension headache | 2 |
| Primary somatoform dizziness | 2 |
| Pseudoradicular cervical syndrome | 2 |
| Recurrent syncope | 2 |
| Retrobulbar neuritis | 2 |

| | |
|---|---|
| Sensitive irritation | 2 |
| Transient monoparesis of the left arm | 2 |
| Trigeminal autonomous headache | 2 |
| vasovagal syncope | 2 |
| Vestibular migraine | 2 |
| gait ataxia | 2 |
| Diffuse hypaesthesia | 3 |
| Dizziness | 3 |
| Episodic tension headache | 3 |
| hemihypaesthesia | 3 |
| hypaesthesia left | 3 |
| Meniere's disease | 3 |
| Myalgia | 3 |
| Orthostatic synkope | 3 |
| Recurrent depressive disorder | 3 |
| Right trigeminal nerve affection | 3 |
| Transient Sensory disturbance | 3 |
| Atypical facial pain | 4 |
| Functional leg paraparesis | 4 |
| Hypertension | 4 |
| Ocular Myasthenia | 4 |
| Headache | 4 |
| Phobic vertigo | 5 |
| Sensory disturbance | 5 |
| Trochlear nerve palsy | 5 |
| Exclusion of chronic inflammatory CNS disease | 6 |
| Trigeminal neuralgia | 6 |
| benign paroxysmal positional vertigo | 7 |
| Unsystematic dizziness | 7 |
| Chronic pain disorder | 8 |
| Tingling paraesthesia | 9 |
| Transient global amnesia | 9 |
| Depression | 14 |
| Dissociative disorder | 16 |
| Migraine | 17 |
| Migraine with aura | 17 |
| Facial palsy | 22 |
| Exclusion of a neuroinflammatory CNS disease | 23 |
| Somatisation disorder | 23 |
| Vestibular neuritis | 32 |
| Tension headache | 75 |

**Z-score values**

Table S2: Age-specific NfL z-scores for 0 and 2 in serum and CSF in control patients:

| Age | Serum z-score 0 [pg/ml] | Serum z-score 2 [pg/ml] | CSF z-score 0 [pg/ml] | CSF z-score 2 [pg/ml] |
|---|---|---|---|---|
| 20 | 5 | 21 | 175 | 776 |
| 25 | 6 | 23 | 200 | 866 |
| 30 | 7 | 26 | 240 | 948 |
| 35 | 8 | 29 | 291 | 1008 |
| 40 | 10 | 32 | 344 | 1044 |
| 45 | 12 | 35 | 398 | 1103 |
| 50 | 15 | 37 | 452 | 1237 |
| 55 | 17 | 41 | 508 | 1485 |
| 60 | 19 | 47 | 578 | 1863 |
| 65 | 22 | 56 | 678 | 2371 |
| 70 | 26 | 70 | 816 | 2971 |
| 75 | 31 | 90 | 995 | 3661 |
| 80 | 36 | 116 | 1192 | 4419 |
| 85 | 43 | 146 | 1400 | 5254 |

Abbreviations: CSF, cerebrospinal fluid; NfL, neurofilament light chain.

### Figures

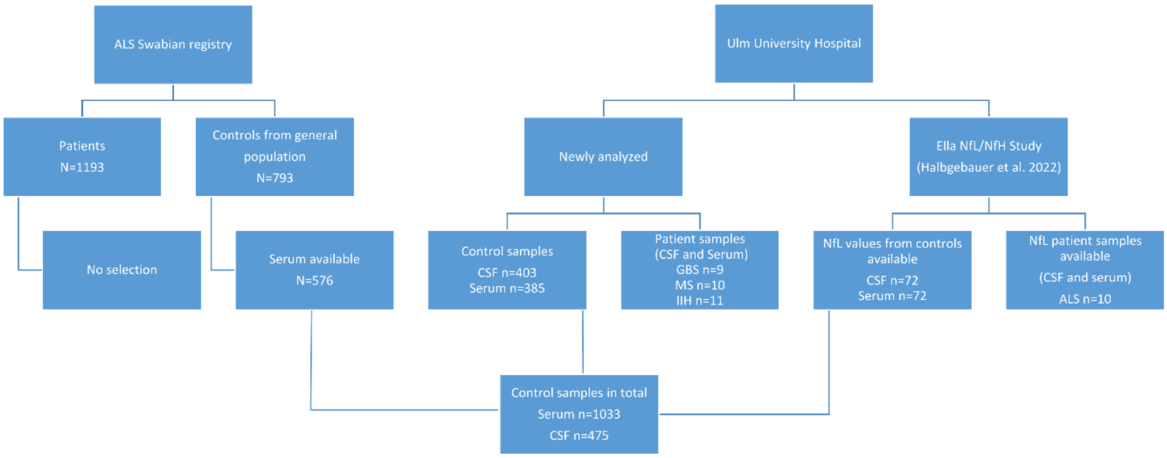


**Figure S1: Patient selection flow chart**

The flow chart describes the selection of controls from the population-based ALS Registry Swabia and the selection of further control patients as well as the ALS, MS GBS and IIH cohorts from patients seen at the University Hospital Ulm. Abbreviations: ALS, amyotrophic lateral sclerosis; Con, control; GBS, Guillain-Barré-Syndrom; IIH, Idiopathic intracranial hypertension; MS, multiple sclerosis; n, number.

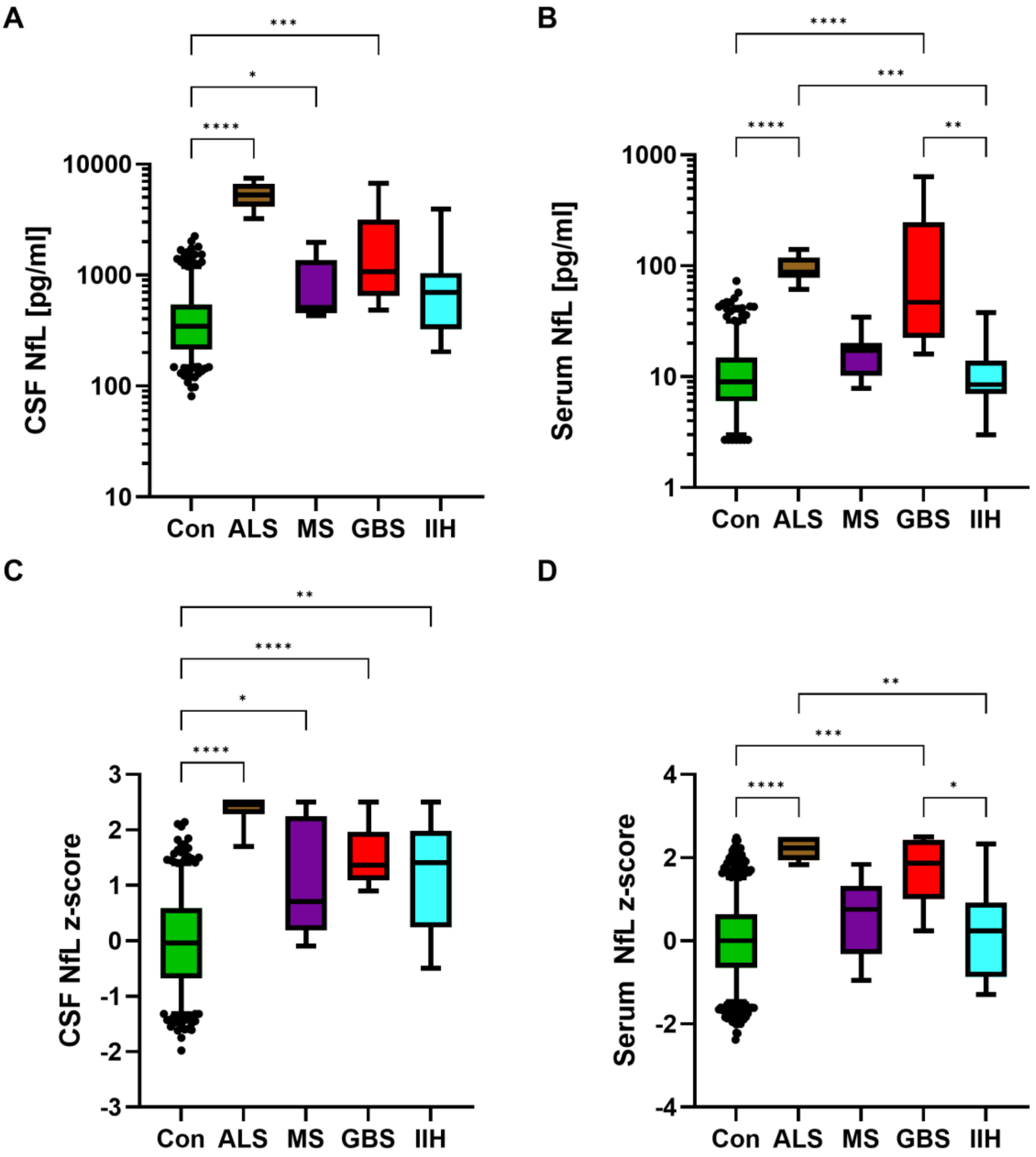


**Figure S2: NfL CSF, serum and Quotient in the disease cohorts**

Figure S1 (**A**) illustrates the CSF NfL absolute values corresponding to the diagnostic group with, as expected, significant higher levels in ALS, MS and GBS. In (**B)** the serum NfL absolute values demonstrate significantly increased concentrations in ALS and GBS. (**C**) displays the CSF NfL z-score values and in (**D**) the z-score values for serum are depicted. *, p<0.05; **, p<0.01; ***, p<0.001; ****, p<0.0001. Abbreviations: ALS, amyotrophic lateral sclerosis; Con, control; GBS, Guillain-Barré-Syndrom; IIH, Idiopathic intracranial hypertension; NfL neurofilament light chain; MS, multiple sclerosis

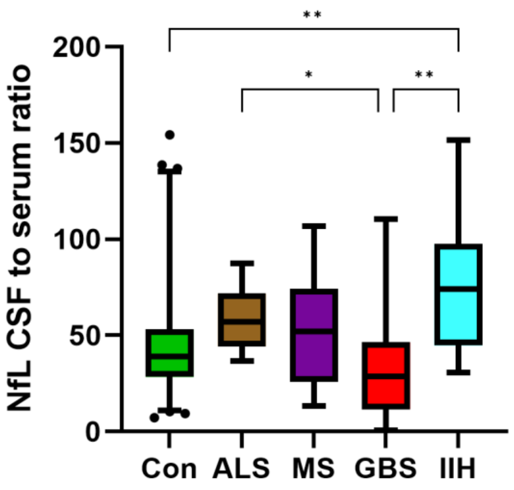


**Figure S3: NfL CSF to serum ratios**

The figure displays the CSF to serum ratio with the highest ratio found in the IIH group.

Abbreviations: Con, control; GBS, Guillain-Barré-Syndrom; IIH, Idiopathic intracranial hypertension; NfL neurofilament light chain

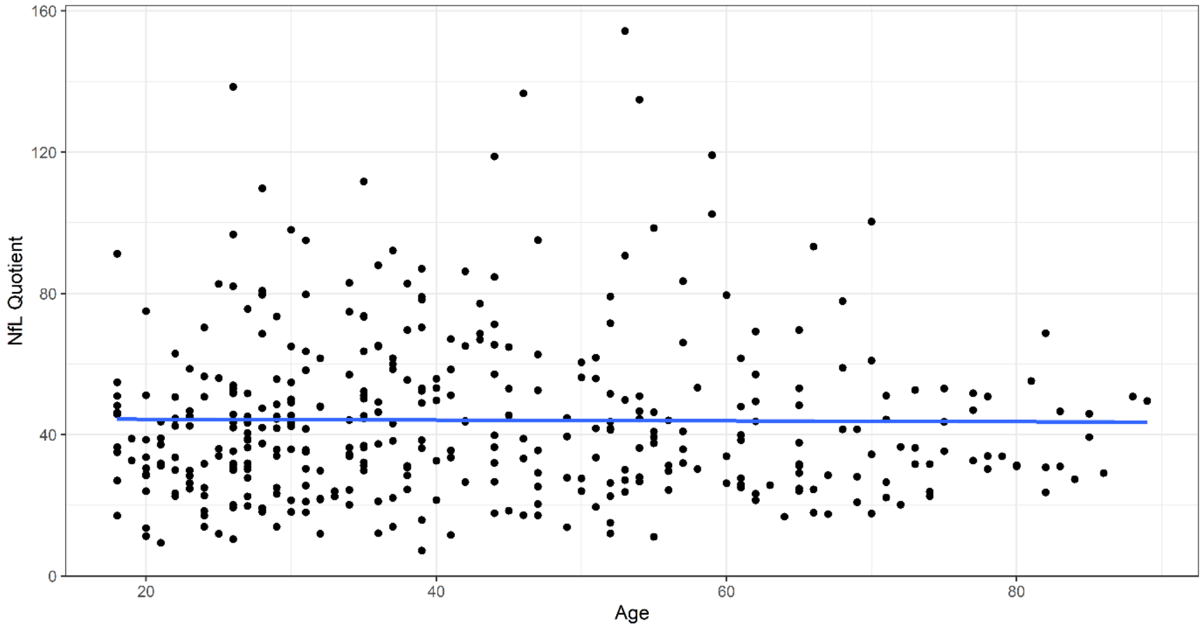


**Figure S4: NfL Quotient and age**

Figure S1 shows the CSF to serum NfL ratio of the control patients corresponding to age. No association with age was detected (r=0.01, -0.1-0.1), p=0.9.Abbreviations: NfL neurofilament light chain

**NfL levels stratified by sex**

Nfl levels in serum and CSF were significantly different between female and male control patients when analyzing the whole cohort (p=0.0001 and p=0.0046, respectively). However, when analyzing NfL levels stratified by sex and age we found no significant difference between female and male controls for serum and CSF (exception: CSF levels in the youngest age group were higher in males) (Figure S5). We therefore didn't create different age-reference graphs for female and male NfL levels.

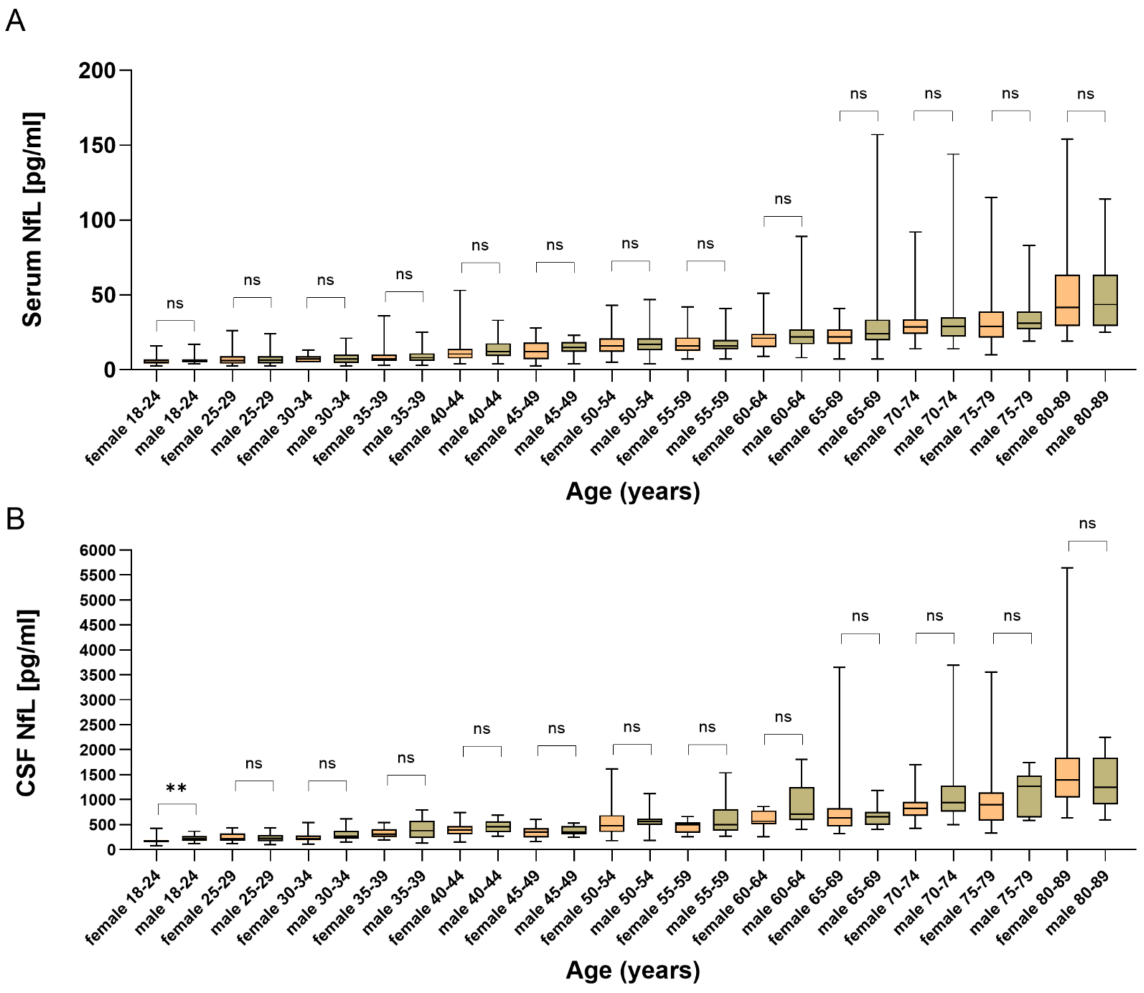


**Figure S5: Serum and CSF NfL in the control cohorts stratified by sex and age**

Figure S2 (**A**) illustrates the serum NfL absolute values corresponding to different age groups stratified by sex. There was no significant difference in any of the age groups (**B)** CSF NfL absolute values stratified by sex and age group. Only the youngest age group depicted significant differences in NfL levels between female and male control patients. there was no difference in all other groups. **, p<0.01; ns, not significant. Abbreviations: CSF, cerebrospinal fluid; NfL neurofilament light chain.

### Influence of the BMI on NfL values

In the literature there is evidence that the BMI has a small effect on NfL concentrations in blood but not CSF [1]. We therefore used the available BMI from 576 controls (age range 45-90 years) to incorporate BMI in the serum NfL model and compare the calculated z-scores to generated z-scores from the model without BMI.

The correlation analysis of NfL and BMI was significant but the spearman r was low (r=-0.14, p<0.0001). Figure S6 depicts the serum NfL values including the BMI as a covariate in the regression model. Table S3 displays the z-scores for the mean BMI (26.58 kg/m$^2$) of the cohort as well as the z-scores for the same cohort without incorporation of the BMI. The data demonstrates that, in our cohort the effect of the BMI on the NfL levels was marginal. Furthermore, we also analyzed the effect of different BMIs (20, 25 and 30 kg/m$^2$) on the NfL levels at certain age groups. Again the difference between NfL levels was small (table S4).

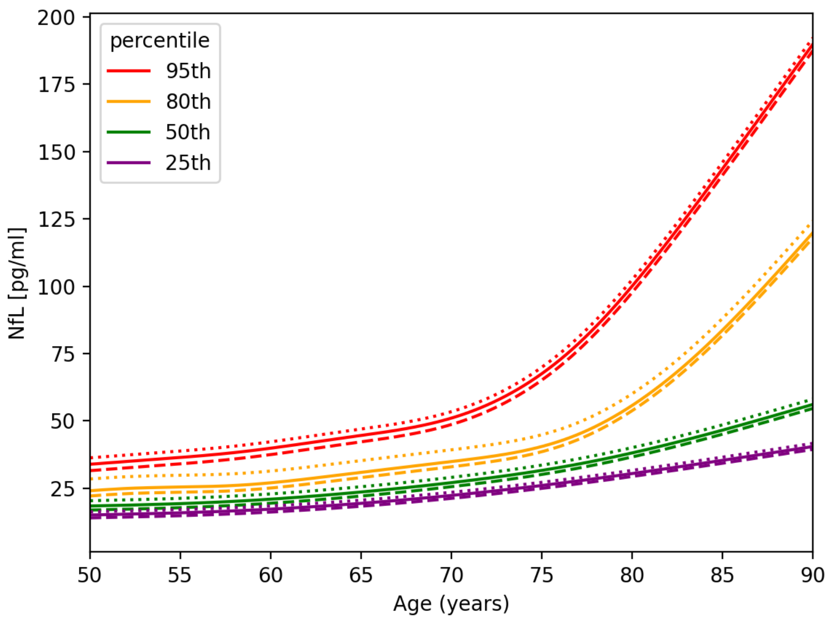


**Figure S6: Serum NfL age-reference curves including BMI in the regression model**
The figure depicts the serum NfL levels as percentiles from 25 to 95 including BMI in the regression model. The solid lines represent an BMI of 25. The dotted lines a BMI of 20 and the dashed lines a BMI of 30. For modelling

the NfL concentration with age and BMI additive quantile regression was applied. Abbreviations: NfL, neurofilament light chain.

Table S3: Comparison of z-scores between the regression model with and without BMI as covariate

| | BMI in the model | w/o BMI in the model | BMI in the model | w/o BMI in the model | BMI in the model | w/o BMI in the model | BMI in the model | w/o BMI in the model |
|---|---|---|---|---|---|---|---|---|
| **Age** | **z=0** | **z=0** | **z=1** | **z=1** | **z=1.5** | **z=1.5** | **z=2** | **z=2** |
| 35 | 14,27 | 15,76 | 24,22 | 22,40 | 30,98 | 29,14 | 38,39 | 37,08 |
| 40 | 15,92 | 16,84 | 23,79 | 23,10 | 29,62 | 28,94 | 38,87 | 37,89 |
| 45 | 17,10 | 17,52 | 22,22 | 23,28 | 28,62 | 29,02 | 39,48 | 38,84 |
| 50 | 17,89 | 18,03 | 24,49 | 25,56 | 30,21 | 31,05 | 41,06 | 40,56 |
| 55 | 18,77 | 18,87 | 26,12 | 27,06 | 32,45 | 33,62 | 43,56 | 43,19 |
| 60 | 20,42 | 20,51 | 27,63 | 28,71 | 35,49 | 36,58 | 48,21 | 47,92 |
| 65 | 23,08 | 23,13 | 31,97 | 32,57 | 40,15 | 40,64 | 56,33 | 56,30 |
| 70 | 26,55 | 26,70 | 36,20 | 36,64 | 45,89 | 46,22 | 68,69 | 69,61 |
| 75 | 31,10 | 31,47 | 42,10 | 43,69 | 57,82 | 57,88 | 89,05 | 90,69 |
| 80 | 37,58 | 38,06 | 59,16 | 60,89 | 80,21 | 79,44 | 119,66 | 120,68 |
| 85 | 46,03 | 46,42 | 88,42 | 88,55 | 110,17 | 108,32 | 157,16 | 156,43 |
| 90 | 55,57 | 55,78 | 123,06 | 121,11 | 142,15 | 139,37 | 196,58 | 193,83 |

Abbreviation; BMI, Body mass index

Table S4: Comparison of z-scores between different BMIs

| | **BMI 20** | **BMI 25** | **BMI 30** | **BMI 20** | **BMI 25** | **BMI 30** | **BMI 20** | **BMI 25** | **BMI 30** | **BMI 20** | **BMI 25** | **BMI 30** |
|---|---|---|---|---|---|---|---|---|---|---|---|---|
| **Age** | **z=0** | **z=0** | **z=0** | **z=1** | **z=1** | **z=1** | **z=1.5** | **z=1.5** | **z=1.5** | **z=2** | **z=2** | **z=2** |
| 35 | 16,78 | 14,82 | 13,32 | 30,04 | 25,25 | 23,09 | 35,32 | 31,80 | 29,77 | 40,51 | 38,89 | 37,28 |
| 40 | 18,44 | 16,48 | 14,98 | 29,61 | 24,82 | 22,66 | 33,96 | 30,45 | 28,41 | 40,98 | 39,37 | 37,75 |
| 45 | 19,62 | 17,66 | 16,16 | 28,04 | 23,25 | 21,09 | 32,96 | 29,45 | 27,41 | 41,60 | 39,99 | 38,37 |
| 50 | 20,40 | 18,44 | 16,95 | 30,31 | 25,52 | 23,36 | 34,55 | 31,03 | 29,00 | 43,17 | 41,56 | 39,94 |
| 55 | 21,29 | 19,33 | 17,83 | 31,94 | 27,14 | 24,98 | 36,79 | 33,28 | 31,24 | 45,67 | 44,06 | 42,45 |
| 60 | 22,93 | 20,98 | 19,48 | 33,45 | 28,66 | 26,50 | 39,83 | 36,31 | 34,28 | 50,32 | 48,71 | 47,10 |
| 65 | 25,60 | 23,64 | 22,14 | 37,79 | 33,00 | 30,84 | 44,48 | 40,97 | 38,93 | 58,45 | 56,83 | 55,22 |
| 70 | 29,07 | 27,11 | 25,61 | 42,02 | 37,22 | 35,07 | 50,23 | 46,72 | 44,68 | 70,80 | 69,19 | 67,58 |
| 75 | 33,61 | 31,65 | 30,15 | 47,92 | 43,13 | 40,97 | 62,16 | 58,64 | 56,61 | 91,16 | 89,55 | 87,93 |
| 80 | 40,10 | 38,14 | 36,64 | 64,98 | 60,18 | 58,02 | 84,55 | 81,03 | 79,00 | 121,77 | 120,16 | 118,54 |
| 85 | 48,55 | 46,59 | 45,09 | 94,24 | 89,44 | 87,29 | 114,51 | 111,00 | 108,96 | 159,27 | 157,66 | 156,04 |
| 90 | 58,09 | 56,13 | 54,63 | 128,88 | 124,08 | 121,93 | 146,49 | 142,97 | 140,94 | 198,69 | 197,08 | 195,46 |

Abbreviation; BMI, Body mass index
